# Correlation of SARS-CoV-2 serology and clinical phenotype amongst hospitalised children in a tertiary children’s hospital in India

**DOI:** 10.1101/2021.01.29.21250660

**Authors:** Aishwarya Venkataraman, S Balasubramanian, Sulochana Putilibai, S Lakshan Raj, Sumanth Amperayani, S Senthilnathan, Anand Manoharan, Arokia Sophi, R Amutha, Kalaimaran Sadasivam, Anu Goenka, A V Ramanan

## Abstract

**Introduction:** Children usually present with minimal or no symptoms of SARS-CoV-2 infection. Antibody responses to SARS-CoV-2 in children from low- and middle-income countries (LMIC) have not been well described. We describe the prevalence of anti-SARS-CoV-2 antibodies and clinical phenotype of seropositive children admitted to a tertiary children’s hospital in South India.

**Methods:** To determine the seropositivity and describe the clinical characteristics of SARS-CoV-2 infection amongst hospitalised children, we performed a prospective clinical data collection and blood sampling of children admitted to Kanchi Kamakoti CHILDS Trust Hospital, Chennai, India over 4 months of the COVID-19 pandemic. In seropositive children, we compared antibody titres between children with and without PIMS-TS.

**Results:** Of 463 children, 91 (19.6%) were seropositive. The median (range) age of seropositive children was 5 years (1 month - 17 years). Clinical presentation was consistent with Paediatric inflammatory multisystem syndrome temporally associated with SARS-CoV-2 infection (PIMS-TS) in 48% (44/91) of seropositive children. The median (range) antibody titre was 54.8 (11.1–170.9) AU/ml among all seropositive children. The median antibody titre among the children with PIMS-TS (60.3 AU/mL) was significantly (p=0.01) higher when compared to the children without PIM-TS (54.8 AU/mL).

**Conclusion:** We describe the antibody responses to SARS-CoV-2 amongst hospitalised children in a LMIC tertiary children’s hospital. Almost half of the seropositive children had PIMS-TS. Antibody levels may be helpful in the diagnosis and disease stratification of PIMS-TS.

**Lay summary:** Children usually present with minimal or no symptoms of SARS-CoV-2 infection. However, Paediatric inflammatory multisystem syndrome temporally associated with SARS-CoV-2 infection (PIMS-TS) has emerged as a distinctive paediatric illness related to SARS-CoV-2. Recently, antibody testing for SARS-CoV-2 is being used increasingly as a diagnostic test for PIMS-TS. However, data on the antibody responses to SARS-CoV-2 in children is sparse. We therefore, attempted to identify the seropositivity and describe the clinical spectrum of SARS-CoV-2 infection amongst infants and children getting hospitalised in a children’s hospital in south India. Nearly one-fifth of the hospitalised children tested serology positive over 4 months. Antibody levels in children with PIMS-TS were significantly higher in comparison to the other two groups (acute SARS-CoV-2 infection and children without PIMS-TS). Results from our study suggest that all children are at risk of SARS-CoV-2 infection though they may present with mild illness or no symptoms. We also observed that antibody testing may have a possible role in diagnosis of PIMS-TS.

## Introduction

Severe acute respiratory syndrome coronavirus 2 (SARS-CoV-2) infection appears to be less severe in children compared with adults.[1–3] Determining the occurrence of SARS-CoV-2 infection in children is an essential facet of understanding the epidemiology of COVID-19 and the possible role of children in transmission.[4–7] Data on SARS-CoV-2 seroprevalence in children has mostly been from developed countries, [5–8] and there are almost no data from low- and middle-income countries (LMIC). We conducted a prospective serological survey to describe the frequency of anti-SARS-CoV-2 antibodies and the associated clinical phenotype in children presenting to a tertiary children’s hospital in Chennai, India.

## Methods

### Study design, setting and participants

We conducted a prospective cross-sectional study of children older than 1 month admitted to Kanchi Kamakoti CHILDS Trust Hospital (KKCTH), a tertiary children’s hospital in Chennai, India from 1 June 2020 to 30 September 2020 and report the symptomatology and clinical findings of infection. The study team approached caregivers of all children admitted to hospital for participation in the study. Demographic, epidemiological and medical data were collected on a standardised case report form. Blood was collected into EDTA tubes (BD Biosciences) and plasma obtained by centrifugation, and frozen at -80°C until use. Study staff involved in serological assays were blinded to clinical data.

### SARS-CoV-2 Antibody assay and RT-PCR test

Antibodies were quantified in plasma using iFlash® SARS-CoV-2 IgG and IgM chemiluminescence antibody assay (CLIA) (YHLO Biotechnology Corporation, Shenzhen, China) according to the manufacturer’s instructions. This is a quantitative CLIA for the detection of IgG and IgM against the SARS-CoV-2 spike (S) and nucleocapsid (N) proteins in human serum/plasma, which has been approved by the Indian Council for Medical Research (ICMR) for SARS-CoV-2 IgG and IgM testing in India[9]. An antibody titre of ≥ 10 AU/ml was considered positive.

Acute COVID 19 and severity of COVID 19 was defined according to the the Ministry of Health and Family Welfare (MOHFW) guidelines [10] issued by Government of India and children with Paediatric inflammatory multisystem syndrome temporally associated with SARS-CoV-2 infection (PIMS-TS) were defined according to the Royal College of Paediatrics and Child Health (RCPCH) case definition for PIMS-TS [11]. SARS-CoV-2 real-time reverse-transcriptase polymerase chain reaction (RT-PCR) performed by Indian Council of Medical Research (ICMR) approved laboratories.

For statistical analysis children were categorised as: Acute COVID 19 (RT-PCR positive) and Serology (IgG) positive. Seropositive children were further categorised as PIMS-TS and Non PIMS-TS Serology positive (IgG).

### Informed consent and ethical approval

Informed consent was obtained from caregivers, and assent was obtained from children where appropriate. The study was approved by the KKCTH CHILDS Trust Medical Research Foundation ethics committee and was registered at Clinical Trials Registry India (CTRI/2020/09/028040).

### Statistical analysis

Continuous variables are presented as medians and interquartile ranges (IQRs), and categorical variables are reported as numbers and proportions. Comparison between the groups was performed using the Mann–Whitney U test and p ≤ 0.05 was considered statistically significant; all tests were two sided. All statistical analyses were performed using SPSS version 24.

## Results

### Basic characteristics

Between June and September 2020, 1311 children were admitted to KKCTH. All the 1311 children were approached for participation, of which 843 were excluded; 102 were less than 1 month old and 741 declined participation. Of the 468 enrolled children, five were excluded due to insufficient blood sample and 463 were included in the final analysis (Figure 1). Children were recruited from the inpatient wards (n=356, 77%), emergency room (n=71, 15%) and paediatric intensive care unit (n=36, 8%). The median age was 5 years old (range 1 month old - 17 years old); and 57% (266/463) were male. SARS-CoV-2 real-time reverse transcriptase polymerase chain reaction (RT-PCR) was positive in 13% (62/463). Anti-SARS-CoV-2 IgG antibodies were detected in 91/463, giving a seropositivity rate of 19.6% (95% CI 15.4 to 22.5, n=463) and the median antibody titre was 54.8 AU/ml (range 11.09 – 170.9). When seropositive children were stratified according to age, 12% (11/91) were under 1 year, 30% (27/11) were 1-5 years old, 46% (42/91) were 5-12 years old and 12% (11/91) were above 12 years old. We did not find any difference in the seropositivity between male and female [20% (54/266) vs 18.7% (37/197), p value = 0.7]. During the 17 week period of the study, the proportion of seropositivity increased which is illustrated in Figure 2.

**Figure 1:**
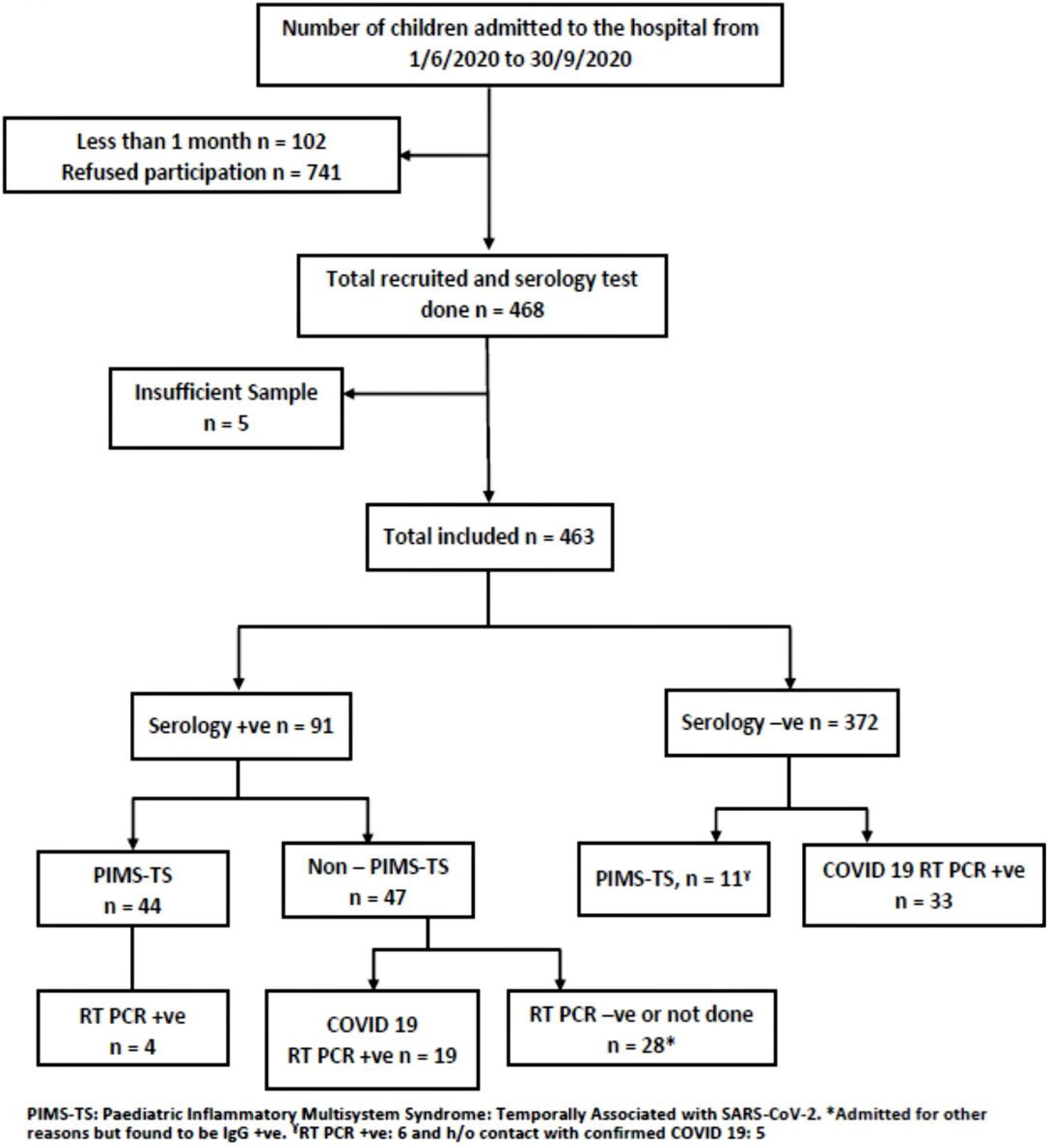
Study overview.

**Figure 2:**
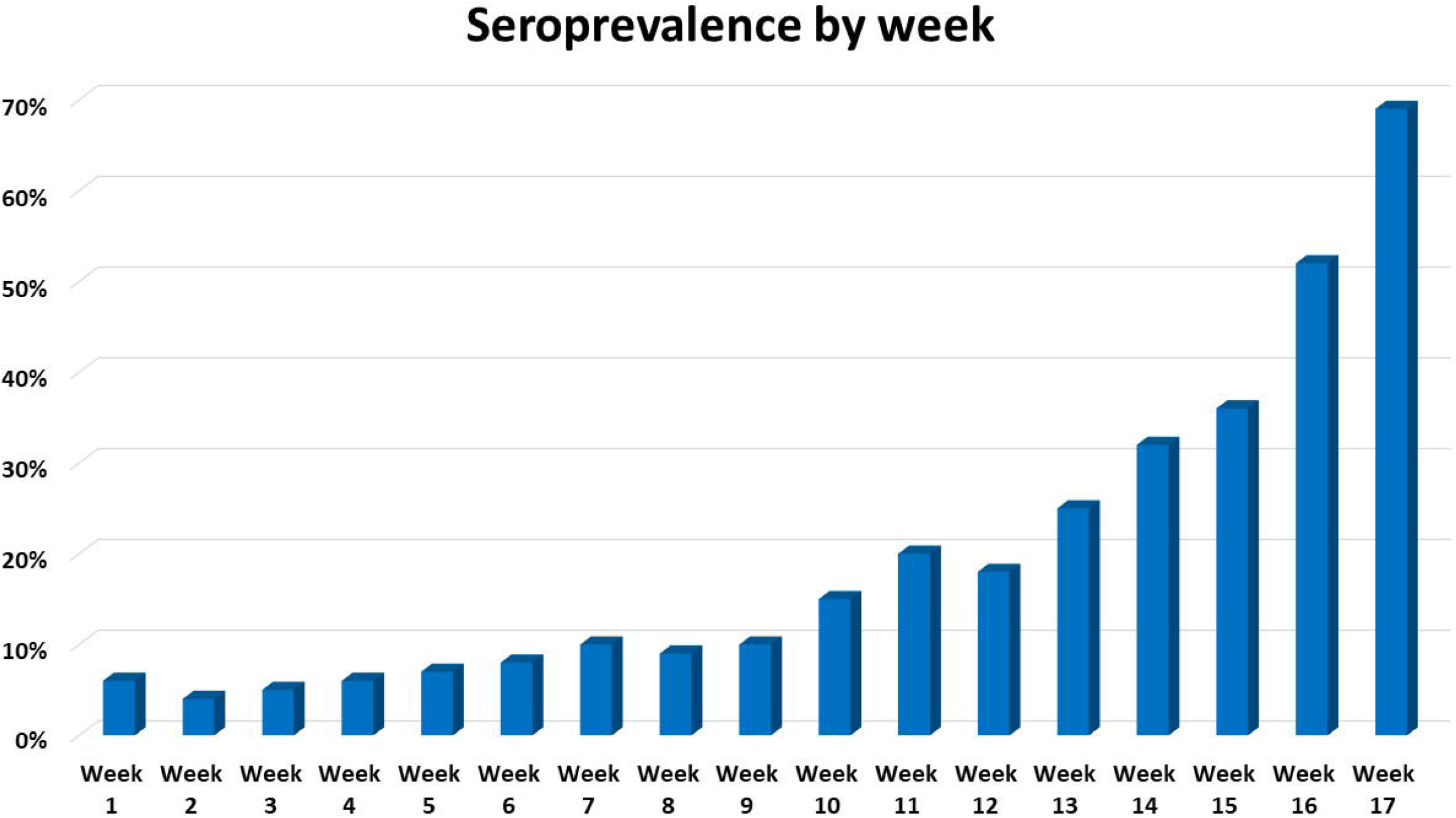
Proportion of seropositive samples obtained each study week from June 1, 2020 to September 30, 2020.

### Characteristics of SARS-CoV-2 (IgG) antibody positive children (Table 1 and Table 2)

Among the 463 children, 19.6% (91) children, with a median age of 5.7 years (range 2 m – 17 y) had a detectable IgG antibody assay, of which 48% (44/91) presented with PIMS-TS. Of the remaining 47 children, 30 children (33%, 30/91) reported no COVID 19 related symptoms. The median antibody titre was 54.8 AU/ml (range 11.09 – 170.9). The most commonly reported symptoms were fever (67%, 61/91) and gastrointestinal symptoms (54%, 49/91). SARS-CoV-2 RT-PCR was positive in 32% (29/91).

A summary of demographics and clinical phenotype of seropositive children is included Table 1. The median antibody titre among the children with PIMS-TS (60.3 AU/mL, range: 12.3 – 170.9) was significantly (p = 0.01) higher when compared to the children without PIM-TS (54.8 AU/ml, range 11.0 – 144.3). In addition to antibody titre, age, gender and clinical phenotype were significantly different between seropositive children with PIMS-TS and children without PIMS-TS (Table 2). There was no significant difference in the median duration since any proven or suspected COVID-19 illness or COVID-19 contact between the PIMS-TS and non-PIMS-TS groups (3 vs 3.2 weeks, p=0.46).

**Table 1:** Characteristics of IgG Serology positive children

|  |  |
| --- | --- |
| **Total IgG Serology +ve (n)** | **91** |
| **Male n (%)** | **54 (59%)** |
| **Age (Median, IQR)** | **5.7 (2 m – 17 y)** |
| **RT-PCR positive n (%)** | **29 (32%)** |
| **Median Antibody titre (IQR) AU/ml** | **54.8 (11.09 – 170.9)** |
| **PIMS-TS n (%)** | **44 (48%)** |
| **Non PIMS-TS n (%)** | **47 (52%)** |
| **Underlying conditions (n=12)** |  |
| *Acute lymphoblastic leukaemia* | **2** |
| *Neurodevelopmental delay* | **1** |
| *Medulloblastoma* | **2** |
| *Inborn error of metabolism* | **2** |
| *Osteosarcoma* | **1** |
| *Atrial septal defect* | **1** |
| *Thalassemia* | **1** |
| *Haemolytic anaemia* | **1** |
| *Seizure disorder* | **1** |
| **Co-existing infection/condition (n=11)** |  |
| *Scrub typhus* | **5** |
| *Dengue viral fever* | **1** |
| *Tuberculosis* | **1** |
| *Urinary tract infection* | **2** |
| *Brucellosis* | **1** |
| *Enteric fever* | **1** |

**Table 2:** Comparison of seropositive positive PIMS-TS and Non PIMS-TS children

|  | **PIMS-TS**<br>**n = 44** | **Non PIMS-TS**<br>**n = 47<sup>#</sup>** | **P value** |
| --- | --- | --- | --- |
| **Age median (years, IQR)** | 7 y (6m – 14y) | 4.4 y (1m – 17 y) | p < 0.05 |
| **Male n (%)** | 19 (43%) | 35 (74%) | p < 0.05 |
| **Median antibody titres (AU/ml)** | 60.3 (12.3 – 170.2) | 54.8 (11.0 – 144.3) | p < 0.05 |
| **RT-PCR positive n (%)** | 10 (23%) | 19 (40%) | p < 0.05 |
| **Underlying conditions n (%)** | 1 (2%) | 11 (23%) | p < 0.05 |
| **Co-existing infections n (%)** | 5 (11%) | 6 (13%) | p = 0.38 |
| **Median duration since proven or suspected COVID illness or contact (weeks, range)** | 3 w (10 d – 4 w) | 3.2 w (10 d – 5 w)<sup>δ</sup> | p = 0.46 |
| **COVID 19 Symptoms n (%)** |  |  |  |
| *Fever* | 44 (100%) | 17 (36%) | p < 0.00001 |
| *Gastrointestinal* | 34 (77%) | 15 (32%) | p < 0.00001 |
| *Respiratory* | 11 (25%) | 16 (34%) | p = 0.17 |
| *Mucocutaneous* | 29 (66%) | 0 | p < 0.00001 |
| *Asymptomatic* | 0 | 30* (64%) | p < 0.00001 |
| **Laboratory parameters** | **n=44** | **n=31** |  |
| *CRP (< 5 mg/L)* | 169 (39 – 473) | 5 (<5 – 181) <sup>¥</sup> | p < 0.00001 |
| *Lymphocyte(/mm<sup>3</sup>) (1500 – 4000) median (IQR)* | 1386 (330 - 2200) | 3890 (650 – 12000) | p < 0.00001 |
| *Neutrophils (/mm<sup>3</sup>) (1500 – 7000) median (IQR)* | 11658 (9918 - 14878) | 6300 (120 – 13160) | p < 0.00001 |
| *Platelets (200 – 450)x10<sup>9</sup>/L median (IQR)* | 110 (62 – 210) | 327 (100 – 540) | p < 0.00001 |
| *Sodium (135 – 145mmol/l) median (IQR)* | 134 (127 -138) | 138 (135 – 150) | p < 0.00001 |
| **Median duration of stay** | 4.5 (3 – 12) | 3 (1 – 10) | p = 0.0001 |
| **Treatment n (%)** |  |  |  |
| *IVIG* | 37 (84%) | 0 | < 0.00001 |
| *Steroids* | 32 (73%) | 2 (4%) | < 0.00001 |
| *PICU admission* | 23 (53%) | 6 (11%) <sup>€</sup> | < 0.00001 |
| *Antibiotics* | 39 (87%) | 16 (31%) | < 0.00001 |
| *Tocilizumab (8mg/kg)* | 2 (4%) | 0 |  |
| **Respiratory support n (%)** |  |  |  |
| *Mechanical Ventilation* | 0 | 0 |  |
| *HHFNC* | 2 (3%) | 2 (4%) |  |
| *Oxygen* | 7 (16%) | 5 (10%) |  |
| **Shock n (%)** |  |  |  |
| *Inotropes* | 23 (52%) | 0 |  |
| *Bolus* | 28 (64%) | 2 (4%) |  |

<sup>#</sup>Including 7 IgM positive. <sup>\*</sup>Including 28 admitted for reasons other than COVID 19, <sup>¥</sup>High CRP seen in children with co-infections, <sup>δ</sup> no history of COVID illness or contact in 4 children, <sup>€</sup>All 6 children were RT-PCR +ve with co-infection. PICU: paediatric intensive care unit; HHFNC: high flow nasal cannula oxygen; IVIG: intravenous immunoglobulin, CRP: C - reactive protein.

### Children with PIMS-TS (Table 3)

A total of 55 children presented with PIMS-TS during the 4 month study duration. Clinical presentation of 19 of these children have been previously described [12]. Of the 55 children, 18% (10/55) had positive SARS-CoV2 RT-PCR test and 80% (44/55) had positive IgG antibody assay. Nearly half (54%, 30/55) of the children with PIMS-TS required PICU admission. The median antibody titre of children with PIMS-TS needing PICU care was significantly lower (45.72 vs 81.28, p = 0.02) in comparison to children who did not require PICU care (Table 3).

**Table 3:** Comparison of children with PIMS-TS needing PICU care and no PICU care

|  | **PIMS-TS admitted to**<br>**PICU n=30** | **PIMS-TS**<br>**No PICU n=25** | **p value**<br>**(PICU vs No PICU)** |
| --- | --- | --- | --- |
| **Age** | 7.2y (11 m – 16 y) | 5y (6 m – 13 y) | 0.116 |
| **Male** | 11 (37%) | 14 (56%) | 0.16 |
| **RT PCR positive** | 8 (26%) | 2 (8%) |  |
| **Serology positive** | 23 (77%) | 21 (84%) | 0.5 |
| **Median antibody titres**<br>**AU/ml** | 45.72 (18.92 – 156.37) | 81.28 (12.32 – 170.21) | 0.02 |
| **Laboratory parameters** |  |  |  |
| **CRP (<5 mg/L)** | 177 (37.5 - 298) | 137 (9 – 370) | 0.34 |
| **D-Dimer (ng/ml FEU)**<br>**(100-500) median (IQR)** | 4890 (2446 – 10000) | 3718 (117 – 10000) | 0.03 |
| **Lymphocyte(/mm<sup>3</sup>)**<br>**(1500 – 4000) median (IQR)** | 1386 (330 – 4540) | 2023 (880 – 6460) | 0.13 |
| **Neutrophils (/mm<sup>3</sup>)**<br>**(1500 – 7000) median (IQR)** | 7800 (3360 – 14180) | 9500 (2700 – 34000) | 0.08 |
| **Sodium (135-145mmol/l)**<br>**median (IQR)** | 133 (130 – 136) | 133 (127 -143) | 0.44 |
| **Ferritin (ng/mL)**<br>**(7 to 140) median (IQR)** | 605 (38 -2571) | 247 (101 – 1104) | 0.006 |
| **LDH (U/L)**<br>**(125-243) median (IQR)** | 494 (200 -905) | 487 (264 -928) | 0.3 |

### SARS-CoV-2 (IgM) antibody positive children

Only 13 children had positive SARS COV2 IgM test, of which 7 had a positive IgG test result and 6 were RT-PCR positive. The median antibody titre was 31.1 AU/ml (range 11. 9 – 139.9). Among the 13 children, 4 were asymptomatic and 6 children had co-exisiting infections (Scrub Typhus, Viral Bronchiolitis, Urinary tract infection). No child had features suggestive of PIMS-TS.

### Characteristics of SARS-CoV-2 RT-PCR positive children (Table 4)

Of the 463 children, 13% (62/463) with a median age of 5 years (4 m – 17 y) tested positive for SARS-CoV2 by RT-PCR; 60% (37/62) were male. A summary of demographic, clinical presentation, investigations, treatment and outcome of children with RT-PCR positive test is shown in Table 4.

**Table 4:** Characteristics of SARS-CoV2 RT-PCR positive children

|  |  |
| --- | --- |
| **Total RT PCR +ve (n)** | **62** |
| **Male n (%)** | **37 (60%)** |
| **Age (Median, IQR)** | **5 (4 m – 17 y)** |
| **Serology Positive** |  |
| **IgG** | **19 (31%)** |
| **IgM** | **6 (9%)** |
| **Median IgG Antibody titre (IQR) AU/ml** | **50.1 (11.09 – 89.0)\*** |
| **COVID 19 Clinical syndrome** | **n (%)** |
| *Mild* | **38 (61%)** |
| *Moderate* | **2 (3%)** |
| *Severe* | **5 (8%)** |
| *Asymptomatic* | **7 (11%)** |
| *PIMS-TS* | **10 (16%)** |
| **Clinical Symptoms** |  |
| *Fever* | **53 (86%)** |
| *Respiratory* | **19 (31%)** |
| *Gastrointestinal* | **14 (23%)** |
| **Underlying conditions (n=8)** |  |
| *Acute lymphoblastic leukaemia* | **2** |
| *Neurodevelopmental delay* | **1** |
| *Nephrotic syndrome* | **2** |
| *Inborn error of metabolism* | **1** |
| *Iron deficiency Anaemia* | **1** |
| *Atrial septal defect* | **1** |
| **Co-existing infection/condition (n=7)** |  |
| *Urinary tract infection* | **2** |
| *Appendicitis* | **1** |
| *Intussusception* | **2** |
| *Brucellosis* | **1** |
| *Tuberculosis* | **1** |
| **Treatment (n=52)‡** |  |
| *Steroids* | **2 (4%)** |
| *PICU* | **6 (11%)** |
| *Antibiotics* | **16 (31%)** |
| *Mechanical Ventilation* | **0** |
| *HHFNC* | **2 (4%)** |
| *Oxygen* | **5 (10%)** |

\*Median titre of 19 IgG positive children. ‡Children with PIMS-TS not included

### Outcome

There were no deaths in our cohort of children.

## Discussion

Studies describing the prevalence of anti SARS-CoV-2 antibodies during the COVID-19 pandemic have been mostly performed in adult populations[4, 13, 14] and paediatric data are lacking particularly from LMIC [5, 7]. To explore the epidemiology of childhood SARS-COV-2 infection in our LMIC setting, we performed SARS-CoV-2 serology on blood samples from children attending a tertiary hospital during a 4-month period of the COVID-19 pandemic in Chennai. We observed a seropositivity of 19.6% in this hospitalised cohort of children, similar to the 21.5% observed in a serosurvey performed by the ICMR among the general population of Chennai during the same period [15, 16]. There was variation in seropositivity among different age groups, however this may not reflect the true paediatric population prevalence because we only sampled children admitted to hospital. It is therefore difficult to infer whether the difference in seropositivity is due to community exposure or varied immunological responses and susceptibility to infection in younger children. Seroprevalence has been reported variably among children[4–8, 17] (<1% to 55%). The seroprevalence in our cohort (19.6%) cannot necessarily be directly compared with those of other studies, given the dissimilarities in population, study-setting, demographics and local SARS-CoV-2 transmission dynamics. Further research is indeed required to understand the differences in infection dynamics between different groups of children as well as to explain the differences across geographic areas.

Over the course of our study, we observed an expected increase in seropositivity among hospitalised children in line with the transmission dynamics of SARS-CoV-2 infection across Chennai during this period.

Almost half of seropositive children in this hospitalised cohort had clinical features consistent with PIMS-TS. This highlights the potentially significant contribution that PIMS-TS plays in driving SARS-CoV-2-related childhood morbidity in LMIC, compared with acute COVID-19 infection. We observed significantly higher antibody titres among seropositive children with PIMS-TS, similar to the other published reports [18, 19]. It remains yet to be understood whether quantitative analysis may have prognostic significance among PIMS-TS children. We found that among the children with PIMS-TS, children needing PICU care had lower titres of IgG antibodies, suggesting a correlation with clinical severity, but this observation requires further study.

We did not find IgM seropositivity commonly among our cohort of children and we observed that IgM antibody status did not correlate with any clinical parameters [20]. In addition, none of the children with PIMS-TS tested positive for IgM antibodies, suggesting that acute COVID 19 and PIMS-TS may have different pathogenesis.

Our study has limited generalisability because it is a single institution study with opportunistic sampling of children attending hospital for medical care for diverse reasons. In addition, antibodies detected in infants may have been transplacentally acquired, though we excluded neonates. We also did not sample children longitudinally, and therefore measurements from seropositive children may have fallen below the limit of detection of the assay due to the time point at which they were sampled.

Despite limitations, our study has strong implications. Firstly, a seropositivity of 19.6% among our cohort of children displays a strong correlation with the general population prevalence. Second, our results suggests that quantitative antibody analysis may be helpful in disease stratification. However, further immunological studies are needed to understand the pathogenesis of SARS-CoV2 infection and disease among children.

## Conclusion

This is the first study describing the SARS-CoV-2 seropositivity among children in India and provides important insight about the SARS-COV-2 infection in children. Our data indicates that children of all age groups in LMIC are at risk of SARS-COV-2 infection. Children exhibit a detectable serological response, and the quantitative analysis may have prognostic significance. SARS-CoV2 IgG antibody levels appear to be higher in children with PIMS-TS.

## Data Availability

The datasets generated during and/or analysed during the current study are available from the corresponding author on reasonable request.

## ACKNOWLEDGEMENTS

We are thankful to Dr Bala Ramachandran, Head paediatric intensive care unit and the paediatric cardiologists Dr Gnanasambandam, Dr Prasad Manne and Dr Kartik Surya for their contribution. We are also thankful to all Paediatric Consultants at Kanchi Kamakoti CHILDS Trust Hospital (KKCTH) for their support and contribution.

## Notes

**CONFLICT OF INTEREST** The Authors have no conflicts of interest to declare

### Competing Interest Statement

The authors have declared no competing interest.

### Clinical Trial

Clinical Trials Registry - India (CTRI) (trial registration: CTRI/2020/09/028040)

### Funding Statement

This study was funded by The CHILDS Trust Medical Research Foundation (CTMRF).

### Author Declarations

The study was approved by the Kanchi Kamakoti Childs Trust Hospital - CHILDS Trust Medical Research Foundation ethics committee

